# Spatial Clustering of Infectious Syphilis in Victoria, Australia: Disentangling MSM Residential Geography from Epidemiological Heterogeneity

**DOI:** 10.64898/2026.02.13.26346302

**Authors:** Hayden Farquhar

## Abstract

**Background:** Victoria, Australia, has experienced a sustained increase in infectious syphilis driven predominantly by transmission among men who have sex with men (MSM). We applied spatial statistical methods to quantify geographic clustering, identify area-level correlates, and disentangle demographic concentration from epidemiological heterogeneity using Census-derived MSM population estimates.

**Methods:** Publicly available notification data for 75 Victorian local government areas (LGAs), 2019–2024, were analysed. Global/Local Moran’s I and Getis-Ord Gi* identified spatial clusters. Spatial regression and geographically weighted regression compared Census same-sex male couple density against a demographic proxy (proportion males aged 20–44). LGA-level MSM populations, estimated using Wilson and Shalley’s Census-based method, were validated against three external benchmarks. Standardised morbidity ratios (SMRs) with Poisson confidence intervals quantified residual per-MSM rate heterogeneity.

**Results:** Strong spatial clustering was identified (Moran’s I = 0.560, p < 0.001), with seven of 11 LISA high-high clusters persisting across all six study years. Same-sex couple density outperformed the demographic proxy (R^2^ = 0.664 vs 0.530; AIC improvement 24.8 units). MSM population adjustment reduced spatial autocorrelation by 74.5% and halved the coefficient of variation of notification rates, though significant residual heterogeneity persisted (Poisson chi-squared p < 0.001). SMR analysis revealed that the highest per-MSM rates were in outer suburban LGAs (Melton 3.25, Frankston 2.06, Brimbank 1.87), while several inner Melbourne hotspot LGAs had SMRs below 1.0. Negative binomial regression with MSM offset identified socioeconomic disadvantage (IRR = 0.995, p < 0.001) and remoteness (IRR = 0.41, p < 0.001) as predictors of per-MSM rates.

**Conclusions:** Three-quarters of Victoria’s apparent syphilis clustering reflects MSM residential geography. Residual per-MSM heterogeneity reveals an outer suburban excess that challenges resource allocation based on raw notification counts and supports decentralised testing expansion.

## Introduction

Victoria, Australia, has experienced a sustained increase in infectious syphilis notifications since approximately 2015, rising from 654 notifications in 2014 to 1,664 in 2023 (rate: 24.1 per 100,000), driven predominantly by transmission among men who have sex with men (MSM) in urban settings.[1,2] Males comprised approximately 80% of notifications, with an estimated 85% of non-Indigenous male notifications nationally attributed to male-to-male sex,[1] though syphilis among females of reproductive age has also increased substantially.[20] Melbourne Sexual Health Centre (MSHC), the state’s principal specialist sexual health service, reported MSM syphilis incidence of 3.7 per 100 person-years (2013–2019; 1,404 cases among 24,391 MSM attendees).[19] Traeger et al. documented national MSM syphilis incidence rising from 2.2 to 4.6 per 100 person-years across ACCESS sentinel surveillance clinics from 2012 to 2022.[2]

Aung et al. described the spatial and temporal epidemiology of infectious syphilis across Victorian LGAs from 2015 to 2018, documenting the generalisation of the epidemic from inner Melbourne to outer suburbs.[3] However, that analysis was descriptive — mapping notification rates without applying formal spatial autocorrelation or regression. A key unanswered question is whether the striking spatial concentration of notifications in inner Melbourne reflects a genuinely more intense epidemic or simply the residential concentration of the at-risk MSM population. Resolving this distinction has direct implications for resource allocation: if the per-MSM epidemic burden is spatially uniform, then areas with lower raw notification rates may have equivalent per-MSM need but inferior case detection.

Outside Australia, formal spatial methods have been applied to syphilis: Salway et al. used spatial regression and GWR in British Columbia, identifying neighbourhood-level structural factors;[5] Tan et al. and Tang et al. applied spatial autocorrelation and regression in China;[6,7] and Schleihauf et al. used spatiotemporal scan statistics in New South Wales.[4] Census-derived same-sex couple data have been used as area-level MSM population proxies in the United States,[14] and Wilson and Shalley developed Australian state-level non-heterosexual population estimates using Census same-sex couple distributions.[21] No Australian study has combined these approaches for syphilis spatial analysis.

This study aims to: (1) quantify the spatial clustering of infectious syphilis across Victorian LGAs using formal spatial statistics; (2) compare area-level demographic and Census-derived MSM proxies as correlates of notification rates; (3) estimate LGA-level MSM populations and validate against external benchmarks; and use (4) standardised morbidity ratios to disentangle demographic concentration from per-MSM epidemiological heterogeneity.

## Methods

### Study design and data sources

This ecological study analysed publicly available aggregate surveillance data. No ethics approval was required as all data were aggregate, de-identified, and publicly accessible.

#### Notification data

Victorian LGA-level infectious syphilis notification counts and estimated resident populations (2019–2024) were obtained from the Victorian Department of Health.[1] Of 79 LGAs in the notification dataset, 75 were linked to the ABS LGA boundary file (ASGS 2021); four (Bayside, Colac-Otway, Kingston, Latrobe) could not be matched due to naming discrepancies. Regression modelling used 74 LGAs (excluding Queenscliffe, which lacked SA2-level covariate data). The mean annual notification rate per 100,000 (2019–2024) was the dependent variable.

#### Demographic data

Population by age and sex (2021 Census, Table G01) was used to calculate the proportion of males aged 20–44 per SA2, aggregated to LGA level using population-weighted spatial joins.[22] Socioeconomic disadvantage was measured using the IRSD.[11] Remoteness classifications were derived from ABS Remoteness Areas.[10]

#### Same-sex couple data

Male-male couple family counts per LGA were obtained from the 2021 Census Same-Sex Couple Indicator (SSCF) via ABS TableBuilder (Counting: Families dataset), and expressed as male-male couples per 1,000 total couple families.[22] This adapts the approach of Grey et al., who used American Community Survey same-sex household data to estimate MSM population sizes at US county level.[14]

#### MSM population estimation

LGA-level MSM populations were estimated following Wilson and Shalley’s Census-based distribution method.[21] Their state-level estimate of 76,267 non-heterosexual males in Victoria (2016, derived from three nationally representative surveys) was scaled to 2021 using male population growth (factor 1.085, yielding approximately 82,750), then distributed to LGAs proportional to each LGA’s share of Victorian male-male couple families. This assumes the geographic distribution of cohabiting same-sex male couples reflects that of the broader MSM population — validated at state level by Wilson and Shalley but untested at LGA level.[21]

#### Service accessibility

Sexual health clinic locations were extracted from the NHSD (AURIN, January 2025).[12] Euclidean distance from SA2 centroids to nearest clinic was aggregated to LGA level.[13]

#### Geographic boundaries

LGA and SA2 boundary files from ABS ASGS 2021.[10] All spatial data used GDA2020 (EPSG:7844).

### Spatial autocorrelation and regression

Queen contiguity spatial weights (mean 5.2 neighbours per LGA) were used for Global/Local Moran’s I,[8] Getis-Ord Gi*,[9] and spatial lag/error regression. Sensitivity was assessed using four alternative weight definitions. Temporal persistence was assessed by computing annual LISA clusters for 2019–2024.

Three regression specifications were compared: (1) demographic proxy (% males 20–44); (2) Census same-sex couple density; and (3) both proxies. All models included log population density, IRSD, log distance to nearest sexual health clinic, and remoteness category. The dependent variable was log(1 + mean annual rate). GWR with adaptive bandwidth assessed spatial non-stationarity.

### MSM-adjusted rate analysis

Estimated LGA-level MSM populations were used to calculate MSM-specific notification rates, validated against three independent external benchmarks: (a) MSHC clinical incidence among MSM (3.7/100 person-years);[19] (b) the proportion of Victorian notifications originating from the inner four LGAs served by MSHC; and (c) national ACCESS sentinel surveillance incidence (4.6/100 person-years in 2022).[2]

Standardised morbidity ratios (SMRs) were calculated as observed/expected notifications, where expected counts were derived from each LGA’s estimated MSM population multiplied by the state-wide MSM notification rate. Exact Poisson 95% confidence intervals were computed. A funnel plot identified LGAs with SMRs significantly outside expected limits. Sensitivity to the MSM population estimate was assessed by varying the Wilson and Shalley scaling factor by ±30%.

Negative binomial regression with log(estimated MSM population) as an offset identified predictors of per-MSM notification rates, providing a count-model complement to the OLS log-rate approach.

### Statistical software

All analyses were conducted in R version 4.5.0 using sf 1.0-19, spdep 1.3-8, spatialreg 1.3-6, spgwr 0.6-37, MASS 7.3-64, tmap 4.1, and tidyverse 2.0.0.[15,16,17]

## Results

### Descriptive epidemiology

A total of 18,096 infectious syphilis notifications were reported across 75 Victorian LGAs during 2019– 2024 (mean 3,016 per year). LGA-level mean annual rates ranged from zero to over 80 per 100,000 in inner Melbourne (Figure 1A), with persistent concentration across the study period (Figure 1B). Hotspot cluster LGAs (n = 11) had higher mean rates (58.6 vs 11.9 per 100,000), higher proportions of males aged 20–44 (21.7% vs 14.1%), and dramatically higher same-sex male couple densities (28.9 vs 4.5 per 1,000 couple families) than non-cluster LGAs (Table 1).

**Table 1.** Characteristics of Victorian LGAs by LISA cluster status, mean 2019–2024.

| Characteristic | Hotspot cluster (n = 11) | Other LGAs (n = 64) |
| --- | --- | --- |
| Mean notification rate per 100,000 | 58.6 | 11.9 |
| Median notification rate per 100,000 | 38.1 | 10.9 |
| Mean % males aged 20–44 | 21.7 | 14.1 |
| Mean SS male couple density (per 1,000) | 28.9 | 4.5 |
| Mean population density (per km<sup>2</sup>) | 3,482 | 308 |
| Mean IRSD score | 1,045 | 993 |
| Mean distance to SH clinic (km) | 6.9 | 106.1 |
| Mean estimated MSM population | 3,343 | 721 |

*LISA = Local Indicators of Spatial Association; SS = same-sex; SH = sexual health; IRSD = Index of Relative Socio-Economic Disadvantage (higher score = less disadvantaged). Source: Victorian Department of Health, ABS Census 2021, NHSD 2025.*

**Figure 1.**
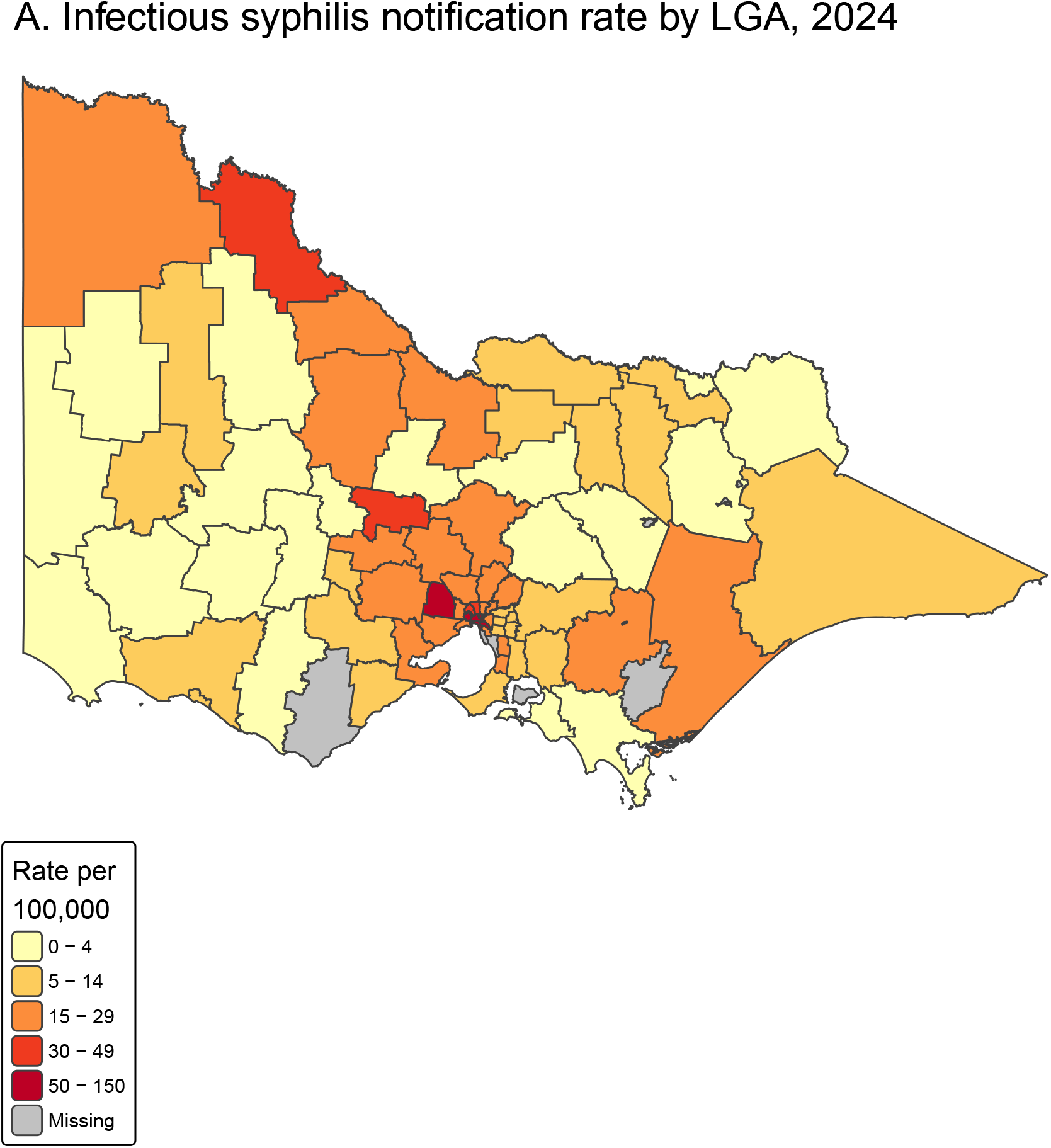
Infectious syphilis notification rates across Victorian local government areas. (A) Mean annual rate per 100,000, 2019–2024. (B) Temporal small multiples for 2019, 2021, and 2024.

### Spatial clustering

Global Moran’s I confirmed significant spatial autocorrelation (I = 0.560, p < 0.001, n = 75), robust across all five weight specifications (I = 0.483–0.573; Supplementary Table S2). LISA identified 11 high-high clusters in inner Melbourne (Figure 2A); Getis-Ord Gi* identified concordant hotspots (Figure 2B).

**Figure 2.**
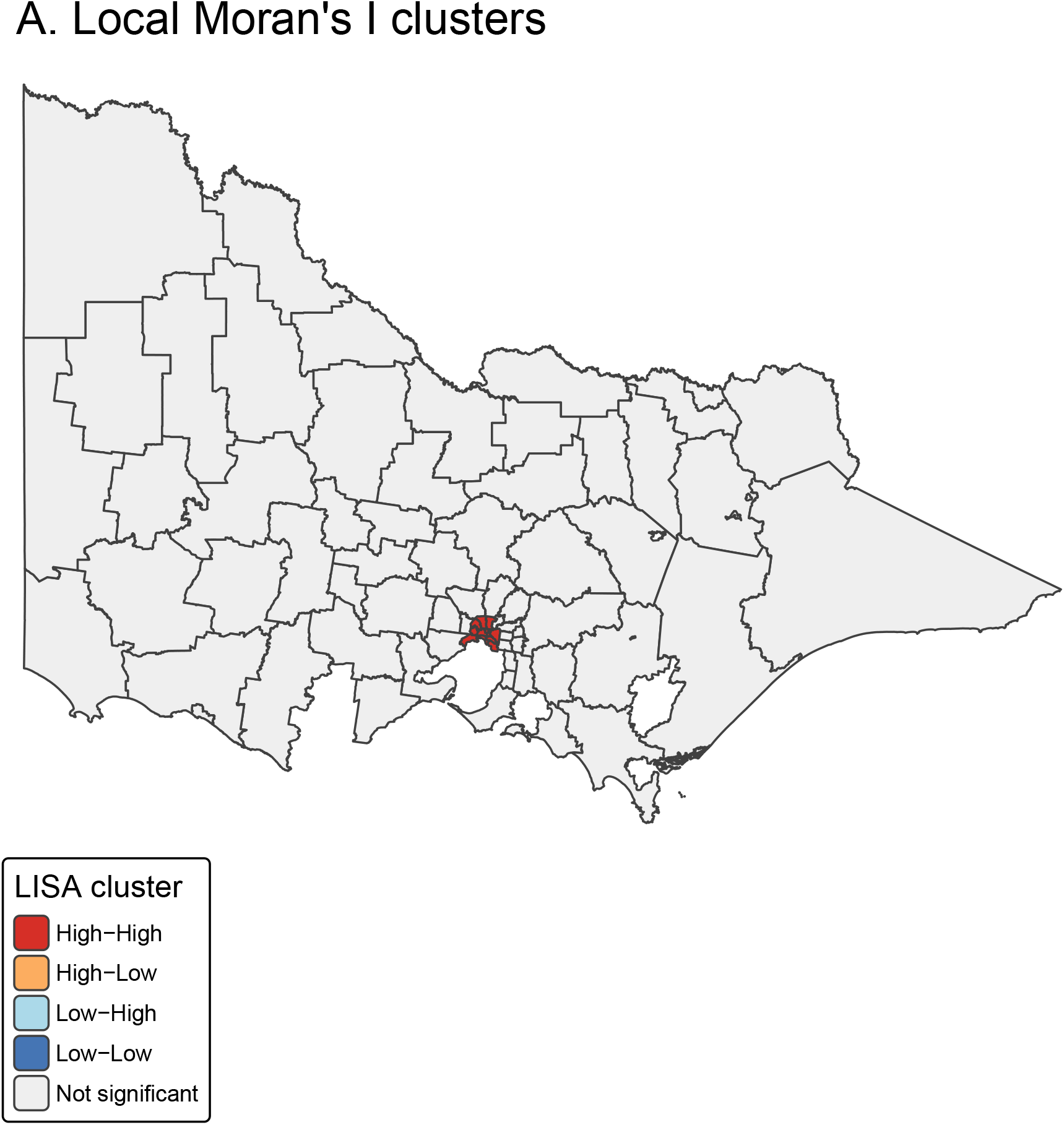
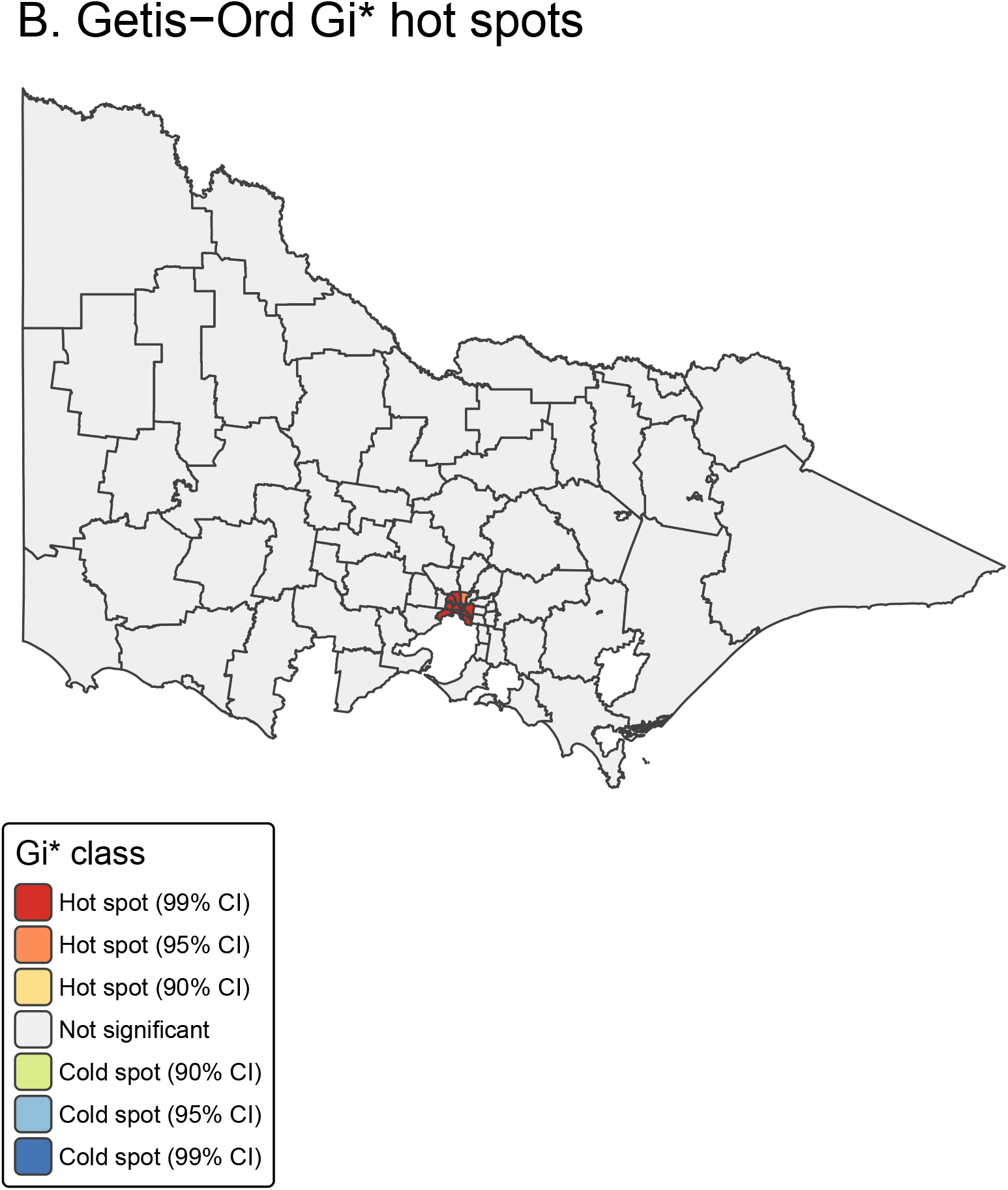
Spatial clustering, Victorian LGAs (n = 75), 2019–2024. (A) LISA cluster classification showing 11 high-high clusters. (B) Getis-Ord Gi* hotspot analysis.

Seven LGAs — Melbourne, Yarra, Port Phillip, Stonnington, Glen Eira, Moreland, and Maribyrnong — were classified as high-high clusters in every year from 2019 to 2024 (Supplementary Table S5), maintaining a ten-fold rate differential over non-cluster LGAs (65–80 vs 5–8 per 100,000; Supplementary Table S6). Moran’s I dipped modestly during 2020–2021 (0.452 vs 0.581 in 2019) but remained significant throughout, consistent with resilience of spatial clustering through Melbourne’s COVID-19 lockdowns.[18] These persistent-cluster LGAs correspond precisely to established MSM community precincts: Commercial Road/Prahran (Stonnington), Fitzroy/Collingwood (Yarra), St Kilda (Port Phillip), the CBD (Melbourne), and Northcote/Brunswick (Moreland) (Supplementary Table S8).

### Census same-sex couple density as MSM proxy

Same-sex male couple density ranged from 2.0 to 49.5 per 1,000 couple families, with Yarra (49.5), Melbourne (41.4), and Port Phillip (34.1) highest. The Pearson correlation between same-sex couple density and the demographic proxy (% males 20–44) was moderate (r = 0.558, p < 0.001), confirming these are related but non-redundant measures. Same-sex couple density correlated more strongly with syphilis notification rates (r = 0.875 vs 0.760).

In spatial regression, the same-sex couple model substantially outperformed the demographic proxy (R^2^ = 0.664 vs 0.530; AIC = 117.9 vs 142.7; Table 2). In the combined model, same-sex couple density remained significant (p < 0.001) while % males 20–44 became non-significant (p = 0.113), confirming that the demographic variable was a noisy proxy for what same-sex couple density captures directly (Supplementary Table S9).

**Table 2.** Spatial regression model comparison: demographic proxy vs Census same-sex couple density, Victorian LGAs (n = 74).

| Variable | Model 1: % Male 20–44 | Model 2: SS couple density |
| --- | --- | --- |
| MSM proxy | 0.110 (0.029)*** | 0.051 (0.007)*** |
| log(Population density) | 0.020 (0.086) | 0.042 (0.070) |
| IRSD score | 0.000 (0.002) | -0.004 (0.002)* |
| log(Distance to SH clinic) | -0.060 (0.151) | -0.020 (0.126) |
| Inner Regional | -0.018 (0.341) | -0.520 (0.300) |
| Outer Regional | -0.163 (0.458) | -0.734 (0.401) |
| **OLS R-squared** | **0.530** | **0.664** |
| **OLS AIC** | **142.7** | **117.9** |
| Lag rho (p-value) | 0.397 (0.002) | 0.214 (0.048) |
| **Lag AIC** | **136.0** | **117.3** |

*OLS coefficients shown as estimate (SE). p<0.05, \*\*p<0.001. Dependent variable: log(1 + mean annual rate per 100,000). IRSD = Index of Relative Socio-Economic Disadvantage. Source: Victorian Department of Health, ABS Census 2021, NHSD 2025.*

**Table 3.** SMR validation: estimated MSM notification rates against external benchmarks.

| Benchmark | Comparator | Our estimate | Ratio | Interpretation |
| --- | --- | --- | --- | --- |
| MSHC incidence [19] | 3.7/100 py | 1.98/100 py (inner 4 LGAs) | 0.53 | 47% below clinic rate — expected |
| Notification share | Inner 4 LGA residential share | 34.9% of VIC total | — | Consistent with MSHC catchment extending beyond 4 LGAs |
| ACCESS<br>incidence [2] | 4.6/100 py<br>(national) | 1.77/100 py (VIC<br>state-wide) | 0.38 | Population-based<br>vs sentinel —<br>expected |

*py = person-years. All comparisons directionally consistent with expected relationships between population-* *based notification rates and sentinel clinic incidence.*

The spatial lag parameter decreased from rho = 0.397 (demographic proxy) to rho = 0.214 (same-sex couple proxy), indicating that approximately half the spatial dependence in the demographic model is consistent with unmeasured MSM residential concentration rather than cross-boundary transmission.

IRSD became significant in the same-sex couple model (OLS p = 0.029; spatial lag p = 0.019), suggesting that socioeconomic disadvantage is associated with higher notification rates once MSM population is properly accounted for — an association masked by demographic confounding in the proxy model.

GWR confirmed spatial non-stationarity (local R^2^ = 0.55–0.84; Figure 3A), with the same-sex couple density coefficient significant in 100% of LGAs.

**Figure 3.**
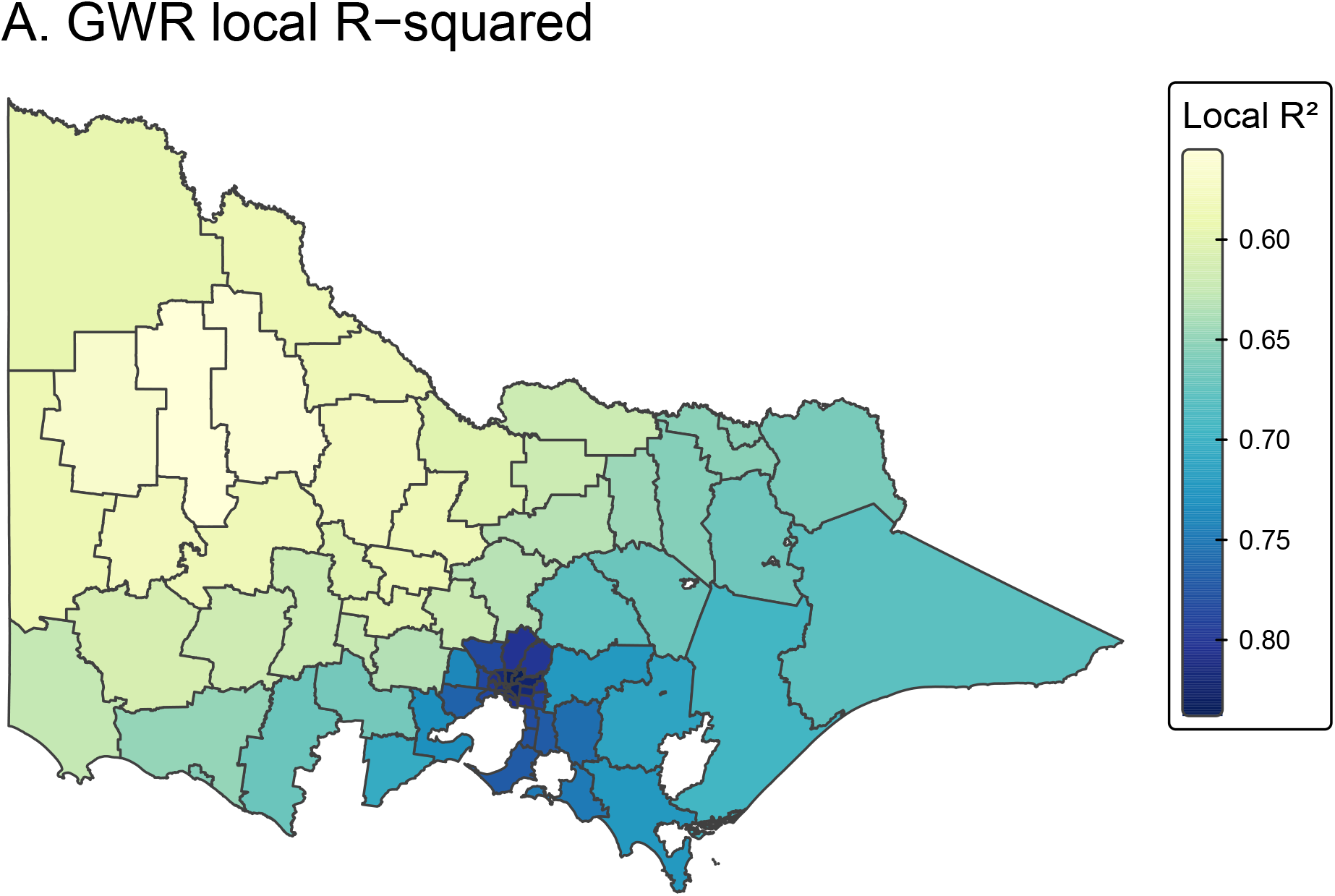
Geographically weighted regression results, Victorian LGAs (n = 74). (A) Local R-squared values (range: 0.55–0.84). (B) Local coefficients for log-transformed distance to nearest sexual health clinic.

### MSM population estimation and validation

Applying Wilson and Shalley’s distribution method yielded estimated MSM populations ranging from approximately 30 (Queenscliffe, West Wimmera) to 8,214 (Melbourne LGA), totalling 82,750 across Victoria. The inner four LGAs (Melbourne, Yarra, Port Phillip, Stonnington) accounted for 31% of the estimated Victorian MSM population.

Three external validation checks supported the plausibility of these estimates:

a. **MSHC benchmark**. The aggregate MSM notification rate for the inner four LGAs was 1.98 per 100 person-years — 47% below the MSHC clinical incidence of 3.7 per 100 person-years.[19] This gap is expected: MSHC attendees are a higher-risk, actively-tested subset, and our denominator includes all estimated MSM regardless of testing behaviour.
b. **Notification share**. The inner four LGAs accounted for 34.9% of all Victorian notifications. Since MSHC (located in Melbourne LGA) draws patients from beyond these four LGAs, the residential share of notifications is expected to be lower than the clinic-diagnosed share, consistent with the observed distribution.
c. **State-wide plausibility**. The Victorian state-wide MSM notification rate was 1.77 per 100 person-years — plausible as a population-based rate against the sentinel clinic incidence of 4.6 per 100 person-years.[2]

### Decomposing apparent clustering: SMR analysis

Adjusting notification rates for estimated MSM population reduced the Global Moran’s I by 74.5% (0.560 to 0.143, p = 0.015), indicating that three-quarters of the spatial autocorrelation in raw notification rates is attributable to the geographic distribution of the MSM population. The coefficient of variation halved from 1.27 (raw) to 0.65 (MSM-adjusted), and MSM residential distribution alone explained 50.6% of the variance in raw notification rates.

The residual Moran’s I remained significant (p = 0.015), and the Poisson homogeneity test formally rejected rate uniformity (chi-squared = 283, df = 74, p < 0.001; overdispersion ratio 3.8), indicating that per-MSM notification rates vary meaningfully across LGAs despite the substantial reduction in spatial heterogeneity. SMR analysis with exact Poisson confidence intervals revealed:

- **14 LGAs with significantly elevated SMRs** (95% CI excludes 1.0)
- **29 LGAs with significantly deficient SMRs**
- **32 LGAs consistent with the state-wide MSM rate**

The highest SMRs were in outer suburban and regional LGAs: Melton (3.25, 95% CI: 2.93–3.60), Swan Hill (2.13), Frankston (2.06), Mildura (1.96), Brimbank (1.87), and Hume (1.69) (Supplementary Table S11; Figure 4). Conversely, several persistent LISA hotspot LGAs had SMRs significantly *below* 1.0: Maribyrnong (0.65), Moreland (0.79), Glen Eira (0.84), and Yarra (0.91) — indicating that their high raw notification rates are entirely explained by their large MSM populations, with per-MSM detection rates actually lower than the state average.

**Figure 4.**
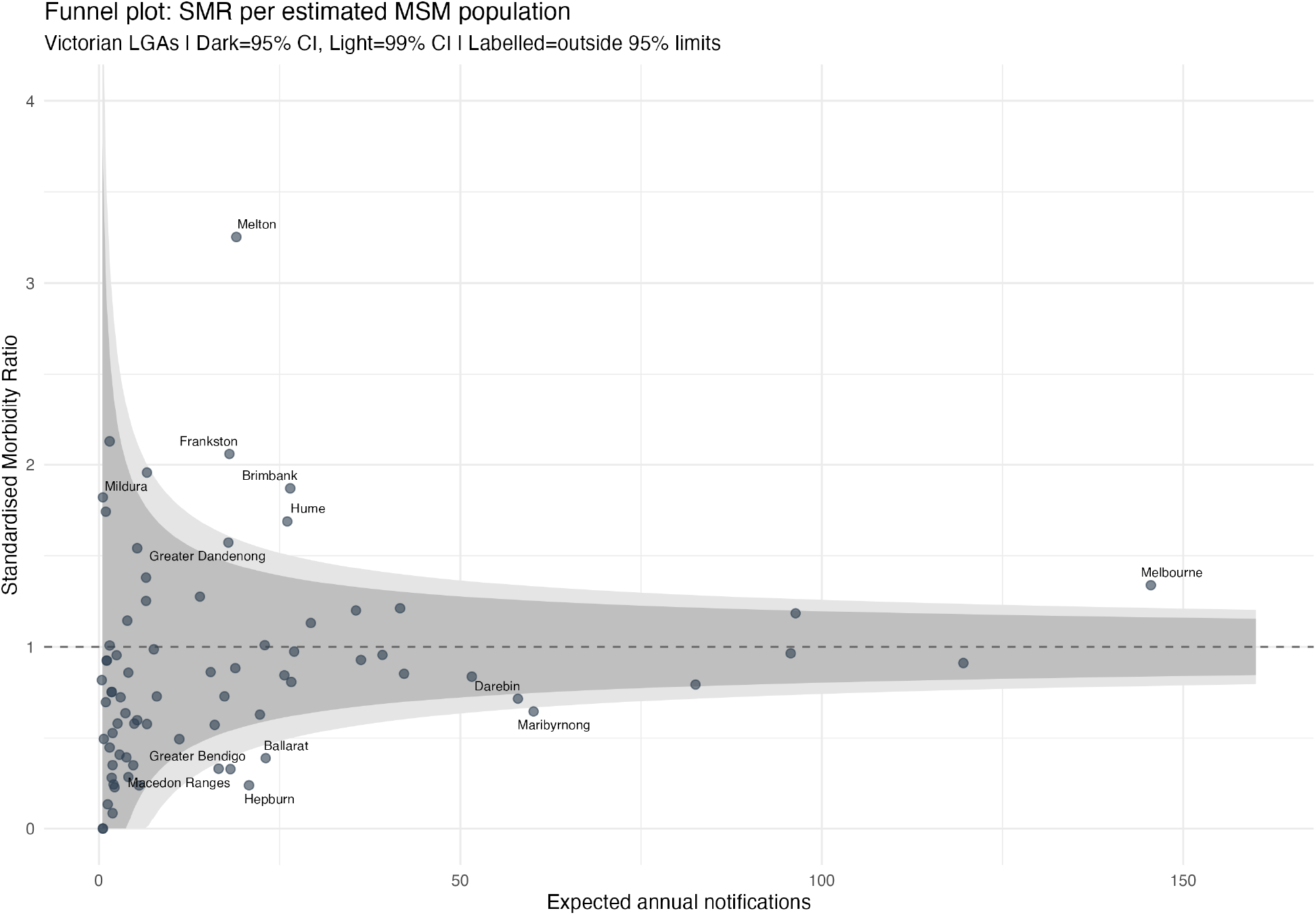
SMR funnel plot: observed/expected syphilis notifications per estimated MSM population, Victorian LGAs. Dark band = 95% Poisson control limits; light band = 99% limits. Note elevated SMRs in outer suburban LGAs (Melton, Frankston, Brimbank) and sub-unity SMRs in some inner Melbourne LGAs (Maribyrnong, Moreland).

Negative binomial regression with MSM population offset (AIC = 394 vs Poisson 444; theta = 14.1) identified two significant predictors of per-MSM notification rates: IRSD score (IRR = 0.995 per unit increase, p < 0.001; lower IRSD [greater disadvantage] associated with higher per-MSM rates) and Inner Regional remoteness (IRR = 0.41, p < 0.001; regional LGAs have approximately 60% lower per-MSM rates). Same-sex couple density was non-significant in the offset model (IRR = 1.001, p = 0.86), confirming that the MSM population is appropriately absorbed by the offset term (Supplementary Table S12).

Spearman correlation of SMRs with area-level characteristics confirmed that population density (rho = 0.324, p = 0.005) and shorter distance to sexual health clinics (rho = -0.241, p = 0.039) were associated with higher per-MSM rates, consistent with a testing intensity gradient. Same-sex couple density was negatively correlated with SMR (rho = -0.265, p = 0.021), indicating that LGAs with the highest MSM concentrations have lower per-MSM notification rates — consistent with mature sexual health infrastructure and higher routine testing coverage in inner Melbourne.

Sensitivity analysis confirmed that the spatial pattern was invariant to the MSM population scaling factor: varying Wilson and Shalley’s estimate by ±30% changed absolute rates but produced identical Moran’s I values and SMR rankings (Supplementary Table S10). Comparison of 2016 and 2021 Census same-sex couple distributions confirmed high temporal stability (Pearson r = 0.983, Spearman rho = 0.972, n = 78 LGAs; Supplementary Figure S8), with all seven persistent hotspot LGAs ranked in the top eight in both Census years, despite a 70% overall increase in male-male couple counts (reflecting marriage equality legislation and increasing disclosure). This stability supports the application of 2021 Census data to the 2019–2024 study period.

## Discussion

This study applied LISA cluster analysis, spatial regression with Census-derived MSM population proxies, and SMR-based decomposition to infectious syphilis notification data across Victorian LGAs — the first application of these combined methods to syphilis data in Australia. The analysis reveals a three-layered spatial structure with distinct policy implications.

### Layer 1: MSM residential geography dominates apparent clustering

Three-quarters of the spatial autocorrelation (Moran’s I reduction from 0.560 to 0.143) and half the total variance in raw notification rates are explained by the geographic distribution of the MSM population alone. Seven inner Melbourne LGAs formed a persistent LISA cluster corridor across all six study years, corresponding precisely to established MSM community precincts.[3] The concordance between formal spatial statistics and known MSM social geography — from Prahran’s Commercial Road to Fitzroy’s Smith Street to St Kilda — provides qualitative validation that the ecological pattern reflects residential demography.

Census same-sex male couple density substantially outperformed the conventional demographic proxy (% males 20–44), achieving an AIC improvement of 24.8 units and a bivariate correlation with syphilis rates of r = 0.875 versus 0.760. This supports the use of Census same-sex couple data for STI spatial analysis in Australia, extending the approach of Grey et al. in the United States[14] and Wilson and Shalley’s population estimation methodology[21] to a public health application. The Pharmaceutical Benefits Scheme prescribing data for HIV pre-exposure prophylaxis (PrEP), available by postcode through the Australian Institute of Health and Welfare, could provide an independent MSM proxy for future validation.

### Layer 2: Residual ascertainment heterogeneity

After MSM population adjustment, residual spatial autocorrelation persisted (Moran’s I = 0.143, p = 0.015). Higher per-MSM notification rates were associated with greater population density and proximity to sexual health clinics — consistent with a testing intensity gradient where areas closer to specialist services detect more cases per MSM.

Socioeconomic disadvantage emerged as a significant predictor of per-MSM rates once MSM population was properly accounted for (NB offset model IRR = 0.995 per IRSD unit, p < 0.001). This association was masked in the standard regression models — demonstrating that proper MSM population adjustment can unmask associations otherwise confounded by demographic composition. The direction of this effect (more disadvantaged areas have higher per-MSM rates) aligns with Salway et al.’s British Columbia findings, where material deprivation was also associated with higher syphilis rates,[5] suggesting a consistent international pattern.

### Layer 3: Outer suburban and regional excess

The LGAs with the highest per-MSM notification rates are not the traditional inner Melbourne hotspots but outer suburban growth corridors: Melton (SMR 3.25), Frankston (2.06), Brimbank (1.87), and Hume (1.69). Meanwhile, several inner Melbourne LGAs — Maribyrnong (0.65), Moreland (0.79), Glen Eira (0.84) — have per-MSM rates below the state average.

This finding should be interpreted with caution given the ecological design and the assumptions underlying the MSM population estimates. Three non-mutually exclusive explanations warrant consideration, with the third carrying particular methodological importance. First, outer suburban LGAs may have genuinely higher per-MSM transmission — consistent with the spatial generalisation of syphilis from inner to outer Melbourne documented by Aung et al. during 2015–2018,[3] which our 2019–2024 data may continue. Second, lower testing access in outer suburbs (which lack specialist sexual health services) may produce diagnostic backlogs, with cases detected later and potentially more advanced. Third, and critically, the Census-derived MSM denominator may be less accurate in outer suburbs: if fewer MSM in these areas are in cohabiting same-sex relationships — due to less accepting social environments, younger demographic profiles, or higher housing costs reducing co-habitation — the Census count would underestimate the true MSM population, systematically inflating the SMR. This third explanation represents an ecological fallacy risk that cannot be resolved with the available data. The sensitivity analysis demonstrates that absolute SMR values change with scaling, but the *relative ranking* of LGAs is invariant; however, differential geographic bias in the cohabiting-to-total MSM ratio would not be captured by uniform scaling. Future studies could address this limitation using Pharmaceutical Benefits Scheme PrEP prescribing data by geography as an independent MSM population proxy, or by obtaining individual-level data on sexual behaviour from sentinel surveillance networks. The high temporal correlation between 2016 and 2021 Census same-sex couple distributions (r = 0.983; Supplementary Figure S8) provides some reassurance that the geographic distribution of the proxy is stable, but does not address whether the cohabiting fraction varies spatially.

Regardless of which explanations dominate, the finding challenges resource allocation based solely on raw notification counts. Inner Melbourne’s high absolute numbers reflect MSM residential concentration, not disproportionate per-MSM burden. Expanding testing infrastructure to outer suburban LGAs could improve population-level case detection.

### Limitations

Several limitations should be noted. First, the ecological design precludes individual-level inference — associations between area-level MSM population and notification rates cannot be attributed to MSM individuals specifically. Second, the MSM population estimation relies on Census same-sex couple data as a geographic distribution proxy.[21] This captures cohabiting couples only (approximately 0.5% of adults in the 2016 Census), missing single, non-cohabiting, and non-disclosing MSM. The geographically constant cohabiting-to-total MSM ratio assumption is untested at LGA level, and differential disclosure between inner urban and outer suburban areas could bias SMR estimates, as discussed above. Third, Victorian LGAs (n = 75) are relatively coarse, and four LGAs (including the populous Kingston [∼170,000] and Bayside [∼110,000]) were excluded due to naming mismatches, potentially affecting LISA cluster detection in south-east Melbourne. Fourth, notification data reflect diagnosed infections; the residual associations with density and clinic distance confirm that ascertainment varies beyond MSM population size alone. Fifth, although the 2021 Census data is applied to a 2019–2024 study period, the high 2016–2021 inter-censal correlation (r = 0.983) and LISA persistence analysis both confirm stability of the MSM residential distribution across this period. Sixth, the log(1 + rate) transformation is an approximation; the negative binomial offset model provides a more appropriate count-data specification and yields consistent conclusions. Only two LGAs recorded zero notifications (estimated MSM populations of approximately 30 each), minimising zero-inflation concerns.

Australia’s syphilis epidemic also includes a heterosexual outbreak disproportionately affecting First Nations communities in northern and central Australia,[1] which this study does not address. The restriction to Victoria — the only jurisdiction providing LGA-level notification data with population denominators — limits generalisability, though the methodology is directly transferable to other states as finer geographic data become available.

## Conclusions

Three-quarters of the spatial autocorrelation in Victorian syphilis notification rates is explained by the residential geography of the MSM population, quantified using Census same-sex couple data. After MSM population adjustment, per-MSM notification rates are substantially more uniform than raw rates but retain meaningful heterogeneity: outer suburban growth corridors (Melton, Frankston, Brimbank) have significantly elevated per-MSM rates while inner Melbourne LGAs have rates at or below the state average.

These findings support four recommendations: (1) resource allocation for syphilis testing should be informed by per-MSM rates rather than raw notification counts; (2) community-based and GP-based testing should be expanded in outer suburban LGAs where the per-MSM burden is highest but specialist infrastructure is absent; (3) syphilis surveillance systems should report estimated MSM-adjusted rates alongside raw rates to enable meaningful geographic comparisons; and (4) the ABS should consider publishing Census same-sex couple data in standard DataPacks at fine geographies to support public health applications.

## Data Availability

All data used in this study are publicly available from the sources described in the Methods section. Analysis code and replication instructions are available at https://doi.org/10.5281/zenodo.19564000.

## Use of artificial intelligence

AI tools (Claude, Anthropic) were used to assist with R code development, data processing pipeline construction, and manuscript drafting and revision. All analyses, scientific interpretations, and methodological decisions were made by the author. The author takes full responsibility for the accuracy and integrity of the work.

## Data Availability

All data used in this study are publicly available from the sources described in the Methods section. Census same-sex couple data were obtained from ABS TableBuilder (free access). Analysis code and replication instructions are available at https://github.com/hayden-farquhar/Syphilis-spatial-analysis (archived: https://doi.org/10.5281/zenodo.19564000).

## Acknowledgements

The following organisations are gratefully acknowledged for making the surveillance and demographic data used in this analysis publicly accessible:

- **Victorian Department of Health** — local government area–level infectious disease notification data (2019–2024)
- **Australian Bureau of Statistics** — Census of Population and Housing 2021 (age-sex structure, same-sex couple indicator, SEIFA), Remoteness Areas, and ASGS boundary files; ABS TableBuilder for Census same-sex couple cross-tabulations
- **Australian Urban Research Infrastructure Network (AURIN)** — access to the National Health Services Directory (NHSD) 2025 snapshot for sexual health clinic locations

## Supplementary material

- **Table S1**. OLS and spatial regression model diagnostics
- **Table S2**. Spatial weight sensitivity
- **Table S3**. GWR local coefficient significance
- **Table S4**. Temporal Moran’s I evolution, 2019–2024
- **Table S5**. LISA cluster persistence by LGA
- **Table S6**. Temporal trends by cluster status
- **Table S7**. Leave-one-out influence analysis
- **Table S8**. MSM precinct validation (100% concordance)
- **Table S9**. Dual-proxy model comparison (3 specifications)
- **Table S10**. MSM population scaling sensitivity (±30%)
- **Table S11**. SMR estimates with Poisson 95% CIs (all 75 LGAs)
- **Table S12**. Negative binomial regression with MSM population offset
- **Figure S1**. OLS residual Q-Q plot
- **Figure S2**. GWR distance significance map
- **Figure S3**. Temporal Moran’s I trend
- **Figure S4**. Rate trends by cluster status
- **Figure S5**. Leave-one-out influence on rho
- **Figure S6**. Proxy validation scatter plot (% Male 20–44 vs SS couple density)
- **Figure S7**. SMR funnel plot (full version with all LGA labels)
- **Figure S8**. Temporal stability: 2016 vs 2021 Census male-male couples by LGA (r = 0.983)

## References

1. Kirby Institute. HIV, viral hepatitis and sexually transmissible infections in Australia: Annual Surveillance Report 2024. Sydney: Kirby Institute, UNSW Sydney; 2024.

2. Traeger MW, Chow EPF, Cornelisse VJ, et al. Syphilis testing, incidence, and reinfection among gay and bisexual men in Australia over a decade spanning HIV PrEP implementation: an analysis of surveillance data from 2012 to 2022. Lancet Reg Health West Pac. 2024;51:101175.

3. Aung ET, Chen MY, Fairley CK, et al. Spatial and temporal epidemiology of infectious syphilis in Victoria, Australia, 2015-2018. Sex Transm Dis. 2021;48(12):e178–e182.

4. Schleihauf E, Watkins RE, Plant AJ. Heterogeneity in the spatial distribution of bacterial sexually transmitted infections. Sex Transm Infect. 2009;85(1):45–49.

5. Salway T, Gesink D, Lukac C, et al. Spatial-temporal epidemiology of the syphilis epidemic in relation to neighborhood-level structural factors in British Columbia, 2005-2016. Sex Transm Dis. 2019;46(9):571–578.

6. Tan NX, Messina JP, Yang LG, et al. A spatial analysis of county-level variation in syphilis and gonorrhea in Guangdong Province, China. PLoS One. 2011;6(5):e19648.

7. Tang S, Shi L, Chen W, et al. Spatiotemporal distribution and sociodemographic and socioeconomic factors associated with primary and secondary syphilis in Guangdong, China, 2005-2017. PLoS Negl Trop Dis. 2021;15(8):e0009621.

8. Anselin L. Local indicators of spatial association — LISA. Geogr Anal. 1995;27(2):93–115.

9. Getis A, Ord JK. The analysis of spatial association by use of distance statistics. Geogr Anal. 1992;24(3):189–206.

10. Australian Bureau of Statistics. Australian Statistical Geography Standard (ASGS) Edition 3, 2021. ABS cat. no. 1270.0.55.001. Canberra: ABS; 2021.

11. Australian Bureau of Statistics. Socio-Economic Indexes for Areas (SEIFA), 2021. ABS cat. no. 2033.0.55.001. Canberra: ABS; 2023.

12. Healthdirect Australia. National Health Services Directory. Accessed via the Australian Urban Research Infrastructure Network (AURIN) open data portal, January 2025.

13. Apparicio P, Abdelmajid M, Riva M, Shearmur R. Comparing alternative approaches to measuring the geographical accessibility of urban health services: distance types and aggregation-error issues. Int J Health Geogr. 2008;7:7.

14. Grey JA, Bernstein KT, Sullivan PS, et al. Estimating the population sizes of men who have sex with men in US states and counties using data from the American Community Survey. Ann Epidemiol. 2016;26(5):304–310.

15. Pebesma E. Simple features for R: standardized support for spatial vector data. R J. 2018;10(1):439–446.

16. Bivand RS, Wong DWS. Comparing implementations of global and local indicators of spatial association. TEST. 2018;27(3):716–748.

17. Tennekes M. tmap: thematic maps in R. J Stat Softw. 2018;84(6):1–39.

18. Chow EPF, Hocking JS, Ong JJ, Phillips TR, Fairley CK. Sexually transmitted infection diagnoses and access to a sexual health service before and after the national lockdown for COVID-19 in Melbourne, Australia. Open Forum Infect Dis. 2020;8(1):ofaa536.

19. Aung ET, Fairley CK, Ong JJ, et al. Incidence and risk factors for early syphilis among men who have sex with men in Australia, 2013-2019: a retrospective cohort study. Open Forum Infect Dis. 2023;10(2):ofad017.

20. Borg SA, Tenneti N, Lee A, Drewett GP, Ivan M, Giles ML. The reemergence of syphilis among females of reproductive age and congenital syphilis in Victoria, Australia, 2010 to 2020: a public health priority. Sex Transm Dis. 2023;50(8):479–484.

21. Wilson T, Shalley F. Estimates of Australia’s non-heterosexual population. Australian Population Studies. 2018;2(1):26–38.

22. Australian Bureau of Statistics. Census of Population and Housing, 2021. Canberra: ABS; 2022.

