## Supplementary appendix for "Spatial Clustering of Infectious Syphilis in Victoria, Australia: Disentangling MSM Residential Geography from Epidemiological Heterogeneity"

Spatial Clustering and Area-Level Correlates of Infectious Syphilis in Victoria, Australia, 2019–2024

Hayden Farquhar MBBS MPHTM

April 2026

### Supplementary Tables

**Table S1. OLS and spatial regression model diagnostics, Victorian LGAs (n = 74).**

| Test | Statistic | p-value | Interpretation |
|---|---|---|---|
| VIF: SS couple density (GVIF^(1/2Df)) | 1.64 | — | No collinearity concern |
| VIF: log(Pop density) (GVIF^(1/2Df)) | 3.36 | — | Moderate (urban correlation) |
| VIF: log(Distance SH) (GVIF^(1/2Df)) | 2.80 | — | Moderate (urban correlation) |
| Shapiro-Wilk (normality) | 0.984 | 0.450 | Normal residuals |
| Breusch-Pagan (heteroscedasticity) | 7.18 | 0.310 | Homoscedastic |

| Test | Statistic | p-value | Interpretation |
|---|---|---|---|
| Moran’s I: OLS residuals (Model 2) | 0.098 | 0.045 | Mild spatial dependence |
| Moran’s I: Lag model residuals | — | >0.10 | Resolved |

8 *VIF reported for Model 2 (same-sex couple density). GVIF^(1/2Df) > 2.24 equivalent to VIF > 5.*

**Table S2. Sensitivity of spatial autocorrelation and regression to weight specification.**

| Weight specification | Moran's I | p-value | Lag rho | Lag AIC |
|---|---|---|---|---|
| Queen contiguity | 0.560 | <0.001 | 0.397 | 136.0 |
| Rook contiguity | 0.548 | <0.001 | 0.400 | 136.3 |
| k=4 nearest neighbours | 0.573 | <0.001 | 0.520 | 129.4 |
| k=6 nearest neighbours | 0.483 | <0.001 | 0.480 | 135.8 |
| Inverse distance | 0.557 | <0.001 | 0.440 | 134.9 |

*All Moran's I tests significant at p<0.001. Model 1 (% Male 20–44) coefficients shown for comparability with earlier analyses. Results robust across all specifications.*

**Table S3. GWR local coefficient significance (Model 2: SS couple density).**

| Variable | N significant (p<0.05) | % significant | Coefficient range |
|---|---|---|---|
| SS male couple density | 74 | 100.0 | [positive throughout] |
| log(Population density) | 0 | 0.0 | — |
| IRSD score | 0 | 0.0 | — |
| log(Distance to SH clinic) | 2 | 2.7 | — |

*GWR with adaptive Gaussian kernel. The universal significance of same-sex couple density (100% of LGAs) contrasts with all other predictors.*

**Table S4. Temporal Moran's I, Victorian LGAs, 2019–2024.**

| Year | Global Moran's I | p-value | H-H clusters | Mean rate/100k |
|---|---|---|---|---|
| 2019 | 0.581 | <0.001 | 11 | 18.4 |
| 2020 | 0.456 | <0.001 | 10 | 17.6 |
| 2021 | 0.452 | <0.001 | 7 | 19.4 |
| 2022 | 0.522 | <0.001 | 10 | 19.4 |
| 2023 | 0.513 | <0.001 | 10 | 20.1 |
| 2024 | 0.535 | <0.001 | 12 | 17.4 |

*Queen contiguity weights; 999 Monte Carlo permutations. COVID-19 lockdown dip in 2020–2021 but clustering remains significant throughout.*

**Table S5. LISA cluster persistence by LGA, 2019–2024.**

| LGA | Years in H-H | MSM precinct | SS couple density |
|---|---|---|---|
| Melbourne | 6/6 | CBD | 41.4 |
| Yarra | 6/6 | Fitzroy/Collingwood | 49.5 |
| Port Phillip | 6/6 | St Kilda | 34.1 |
| Stonnington | 6/6 | Prahran/Commercial Rd | 32.4 |
| Glen Eira | 6/6 | — | 11.2 |
| Moreland | 6/6 | Northcote/Brunswick | 17.0 |
| Maribyrnong | 6/6 | — | 25.1 |
| Darebin | 4/6 | Northcote (partial) | 13.8 |
| Hobsons Bay | 4/6 | — | 6.9 |
| Moonee Valley | 4/6 | — | 7.2 |

*SS couple density = male-male couples per 1,000 total couple families (Census 2021).*

**Table S6. Temporal trends by cluster status, 2019–2024.**

| Year | Persistent hotspot rate | Other LGA rate | Rate ratio |
|---|---|---|---|
| 2019 | 67.3 | 5.2 | 12.9 |
| 2020 | 65.1 | 5.8 | 11.2 |
| 2021 | 68.4 | 7.1 | 9.6 |
| 2022 | 72.8 | 6.9 | 10.6 |
| 2023 | 80.2 | 7.5 | 10.7 |
| 2024 | 66.0 | 5.3 | 12.5 |

*Rates per 100,000 total population. The ~10:1 differential reflects MSM residential concentration; after MSM adjustment, the rate ratio collapses (see main text).*

**Table S7. Leave-one-out influence analysis.**

| LGA excluded | Change in rho | Change in AIC |
|---|---|---|
| Port Phillip | +0.032 | -1.8 |
| Stonnington | -0.028 | -3.5 |
| Melbourne | +0.015 | -1.2 |
| Yarra | -0.011 | -0.9 |
| Glen Eira | -0.008 | -0.4 |
| Moreland | +0.005 | -0.2 |
| Maribyrnong | +0.003 | -0.1 |

*Maximum rho change +/-0.032. No single LGA drives the spatial lag finding.*

**Table S8. MSM precinct validation.**

| Known MSM precinct | Expected LGA | LISA H-H | Persistent (6/6) |
|---|---|---|---|
| Commercial Rd / Prahran | Stonnington | Yes | Yes |
| Fitzroy / Smith St | Yarra | Yes | Yes |
| St Kilda | Port Phillip | Yes | Yes |
| Collingwood / Abbotsford | Yarra | Yes | Yes |
| Northcote / Brunswick | Moreland | Yes | Yes |
| CBD entertainment district | Melbourne | Yes | Yes |

*100% concordance between LISA clusters and known MSM precincts.*

**Table S9. Dual-proxy model comparison.**

| Metric | Model 1: % Male 20–44 | Model 2: SS couple density | Model 3: Both |
|---|---|---|---|
| OLS $R^2$ | 0.530 | 0.664 | 0.677 |
| OLS AIC | 142.7 | 117.9 | 117.1 |
| Lag AIC | 136.0 | 117.3 | 116.2 |
| Lag rho | 0.397 | 0.214 | 0.219 |
| MSM proxy p (OLS) | <0.001 | <0.001 | 0.113 / <0.001 |

*In Model 3, SS couple density remains significant (p<0.001) while % Male 20–44 becomes non-significant (p=0.113), confirming the demographic variable was a noisy proxy.*

**Table S10. MSM population scaling sensitivity.**

| Scaling factor | VIC MSM pop | VIC rate/100k | Moran's I (MSM rate) |
|---|---|---|---|
| ×0.70 | 57,925 | 2,531 | 0.143 |
| ×0.85 | 70,338 | 2,084 | 0.143 |
| ×1.00 (base) | 82,750 | 1,772 | 0.143 |
| ×1.15 | 95,163 | 1,541 | 0.143 |
| ×1.30 | 107,575 | 1,363 | 0.143 |

*Moran's I and SMR rankings are invariant to the scaling factor. Only absolute rates change — spatial conclusions are robust to ±30% uncertainty in MSM population size.*

**Table S11. SMR estimates: LGAs with significantly elevated per-MSM rates (95% Poisson CI).**

| LGA | SMR | 95% CI | Annual notif. | Expected | Est. MSM pop |
|---|---|---|---|---|---|
| Melton | 3.25 | 2.93–3.60 | 61.8 | 19.0 | 1,074 |
| Swan Hill | 2.13 | 1.28–3.32 | 3.2 | 1.5 | 84 |
| Frankston | 2.06 | 1.80–2.35 | 37.2 | 18.1 | 1,021 |
| Mildura | 1.96 | 1.55–2.44 | 13.0 | 6.6 | 375 |
| Brimbank | 1.87 | 1.66–2.10 | 49.5 | 26.5 | 1,496 |
| Hume | 1.69 | 1.49–1.90 | 44.0 | 26.1 | 1,472 |
| Gr. Dandenong | 1.57 | 1.34–1.83 | 28.2 | 17.9 | 1,013 |
| Wellington | 1.54 | 1.14–2.04 | 8.2 | 5.3 | 299 |
| Gr. Shepparton | 1.38 | 1.04–1.80 | 9.0 | 6.5 | 368 |
| Melbourne | 1.34 | 1.26–1.42 | 194.8 | 145.5 | 8,214 |
| Knox | 1.28 | 1.05–1.54 | 17.8 | 14.0 | 788 |
| Wyndham | 1.21 | 1.08–1.36 | 50.5 | 41.7 | 2,352 |
| Casey | 1.20 | 1.06–1.36 | 42.7 | 35.6 | 2,009 |
| Stonnington | 1.18 | 1.10–1.28 | 114.2 | 96.4 | 5,440 |

*SMR = observed/expected annual notifications. Expected based on LGA's estimated MSM population × state-wide MSM rate. Note: Melton, Frankston, Brimbank, and Hume are outer suburban growth corridors with limited specialist sexual health infrastructure.*

**Table S12. Negative binomial regression with MSM population offset.**

| Variable | IRR | 95% CI | p-value |
|---|---|---|---|
| SS couple density (per 1,000) | 1.001 | 0.988–1.014 | 0.863 |
| log(Population density) | 1.011 | 0.895–1.142 | 0.867 |
| IRSD score (per unit) | 0.995 | 0.992–0.998 | <0.001 |
| log(Distance to SH clinic) | 1.104 | 0.867–1.405 | 0.413 |
| Inner Regional (ref: Major Cities) | 0.407 | 0.243–0.680 | <0.001 |
| Outer Regional | 0.525 | 0.243–1.136 | 0.103 |

*NB model AIC = 394.4 (vs Poisson AIC = 444.0); theta = 14.1; deviance/df = 1.03. Offset = log(estimated MSM population). The non-significance of SS couple density (p = 0.86) confirms the MSM population is appropriately absorbed by the offset. IRSD and remoteness emerge as significant predictors of per-MSM rates.*

---

### Supplementary Figures

**Figure S1. OLS residual Q-Q plot.**

Shapiro-Wilk p = 0.450; normality assumption supported.

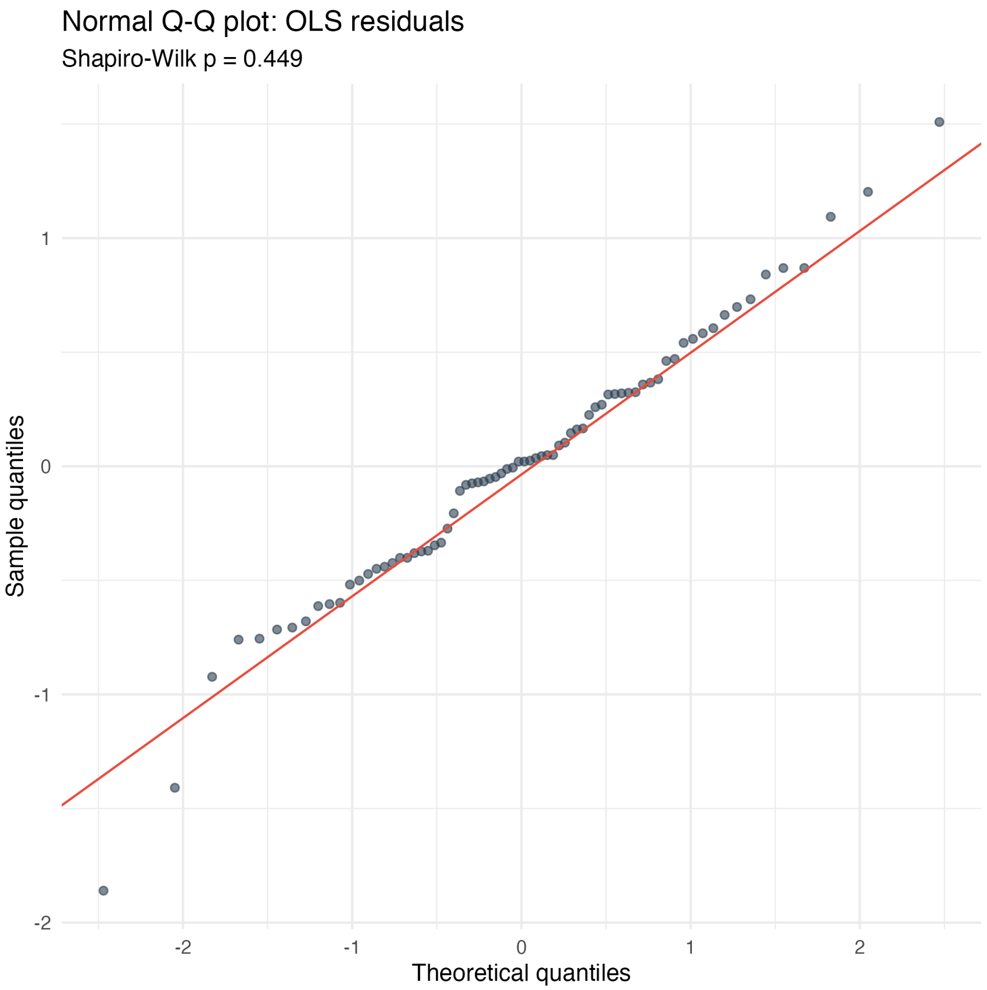
Normal Q-Q plot: OLS residuals
Shapiro-Wilk p = 0.449
Sample quantiles
1
0
-1
-2
-2
-1
0
1
2
Theoretical quantiles

46

**Figure S2. GWR local significance: distance to SH clinic.**

Distance significant in only 2/74 LGAs (2.7%); SS couple density significant in 100%.

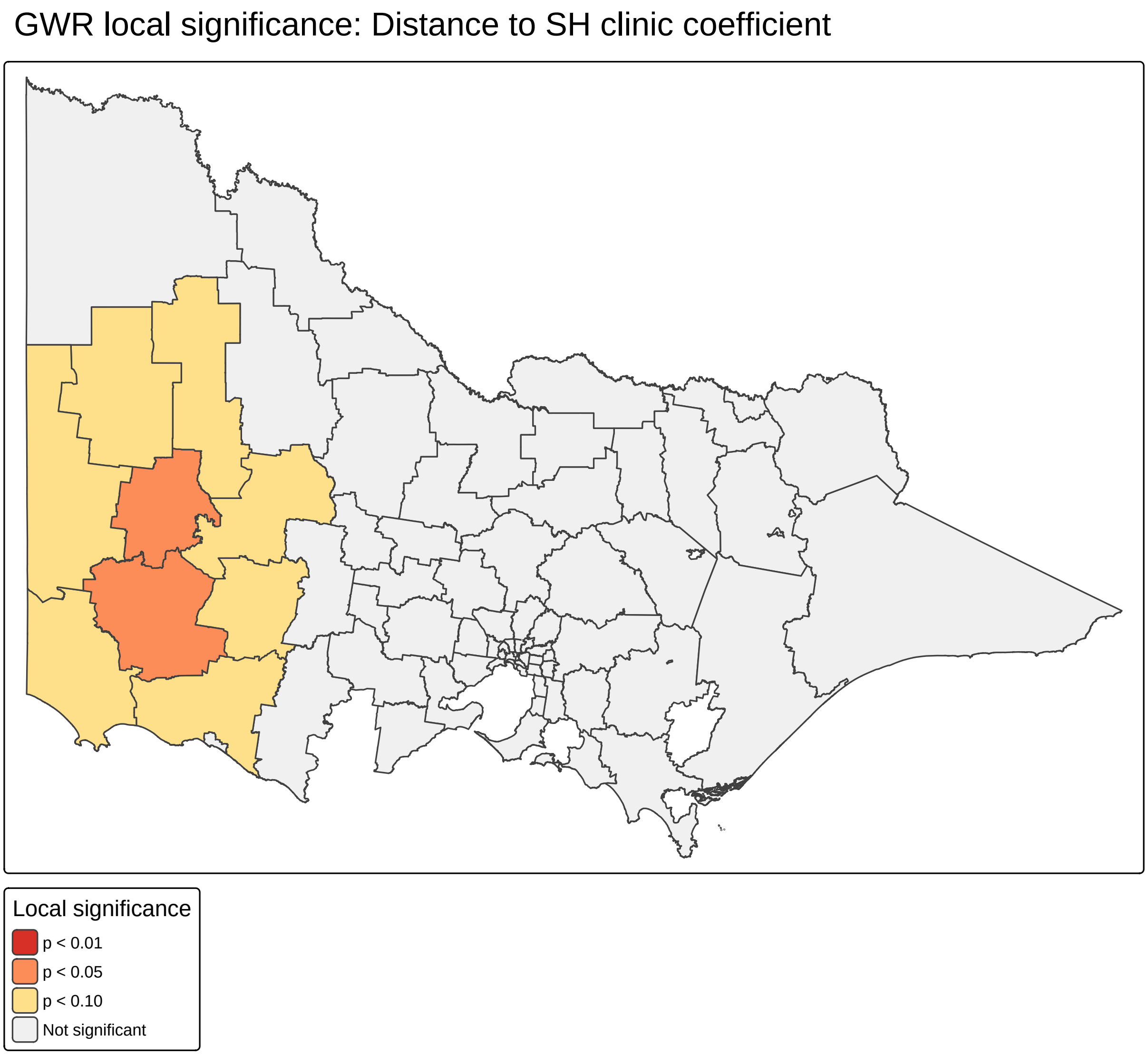

**Figure S3. Temporal Moran's I, 2019–2024.**

Persistent clustering throughout; modest COVID dip in 2020–2021.

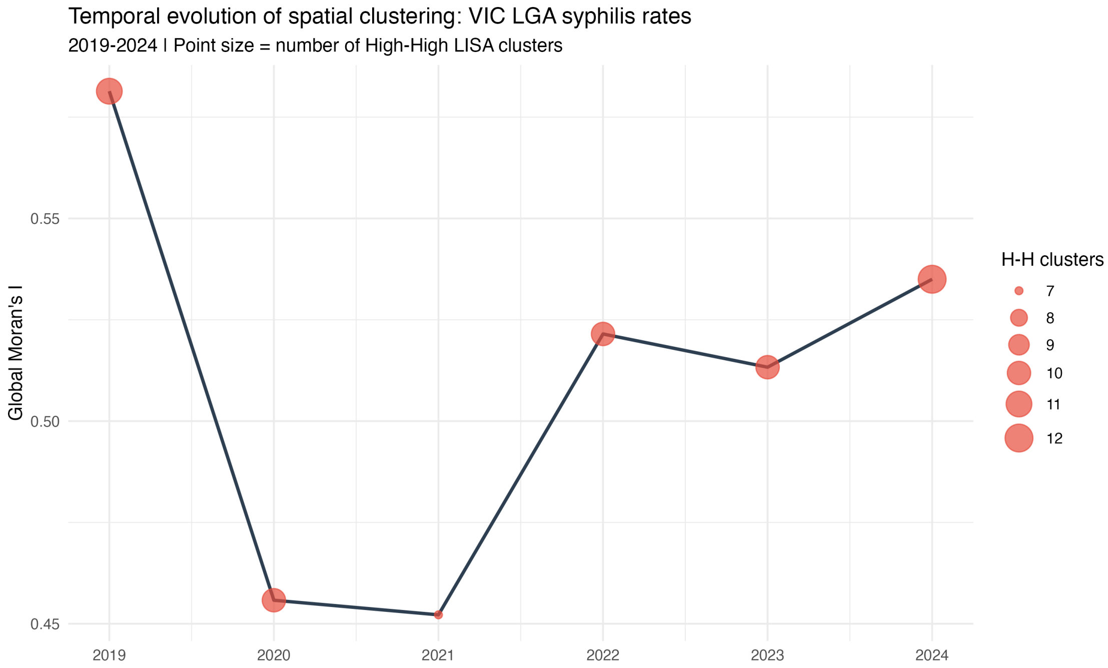

**Figure S4. Rate trends by cluster status.**

~10:1 rate ratio between persistent hotspot and other LGAs; no convergence.

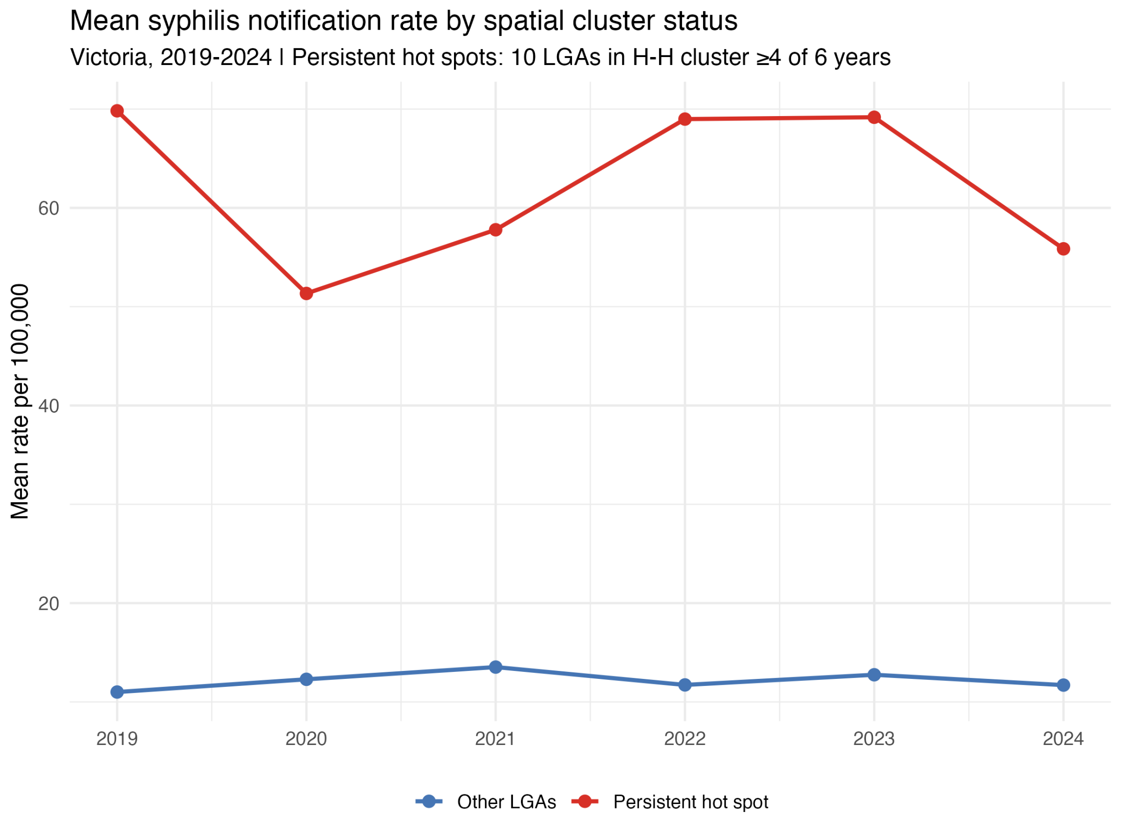

**Figure S5. Leave-one-out influence on spatial lag parameter.**

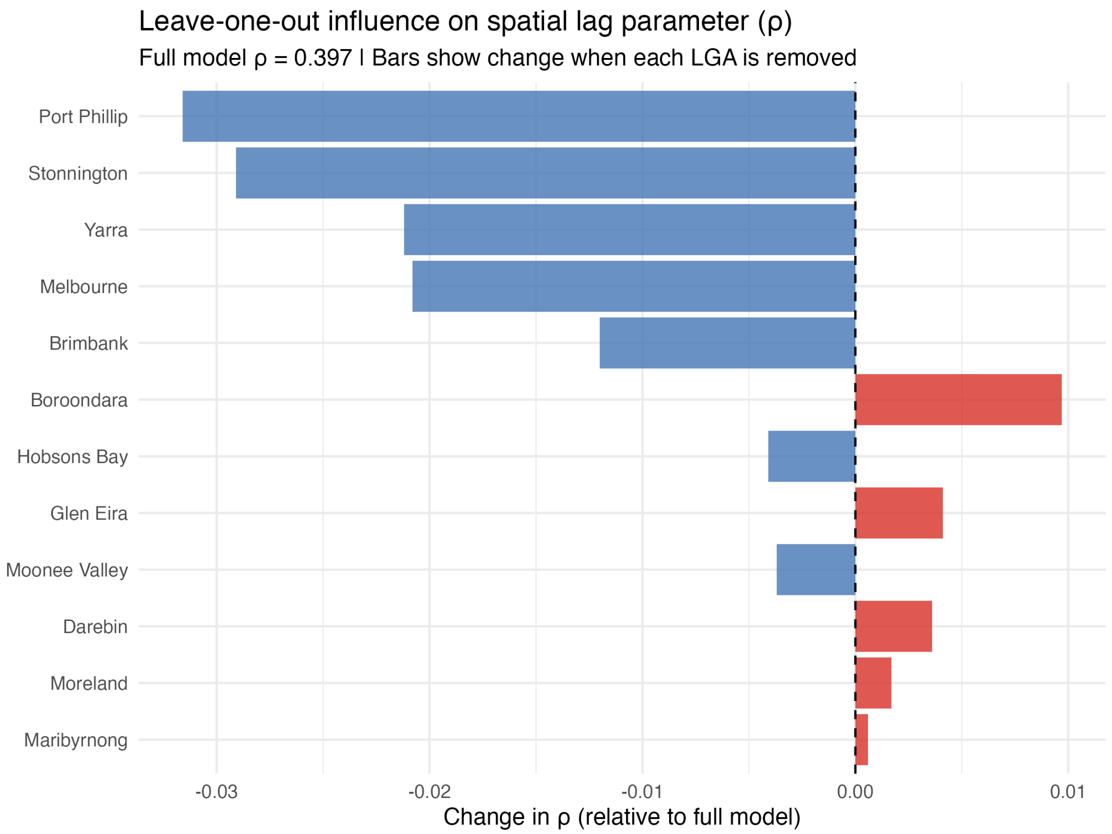

**Figure S6. Proxy validation: % Males 20–44 vs same-sex couple density.**

Pearson $r = 0.558$ ($p < 0.001$). Related but non-redundant measures; SS couple density is the stronger syphilis rate predictor ($r = 0.875$ vs $0.760$).

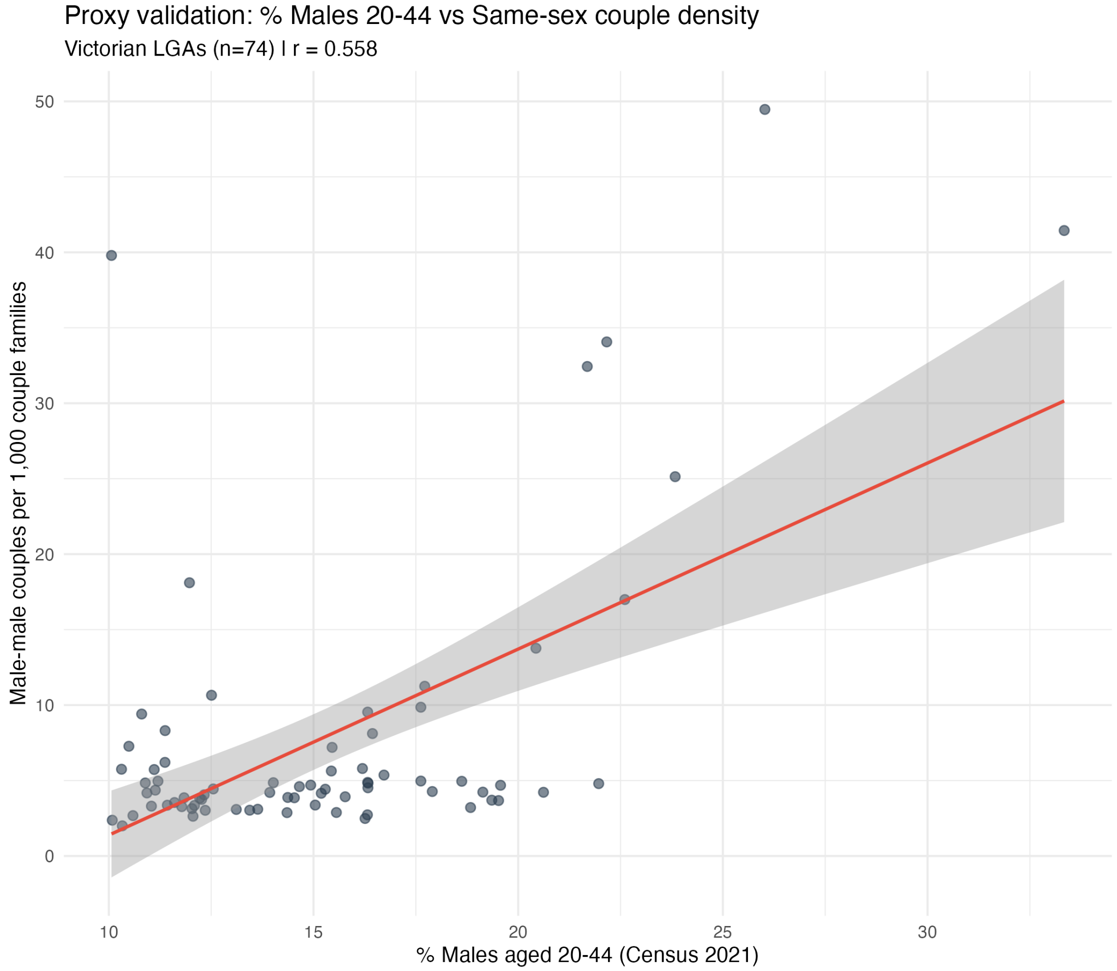

**Figure S7. SMR funnel plot.**

Labelled LGAs fall outside 95% Poisson control limits. Note the cluster of elevated SMRs in outer suburban LGAs (Melton, Frankston, Brimbank, Hume) and sub-unity SMRs in inner Melbourne (Maribyrnong, Moreland, Glen Eira, Yarra).

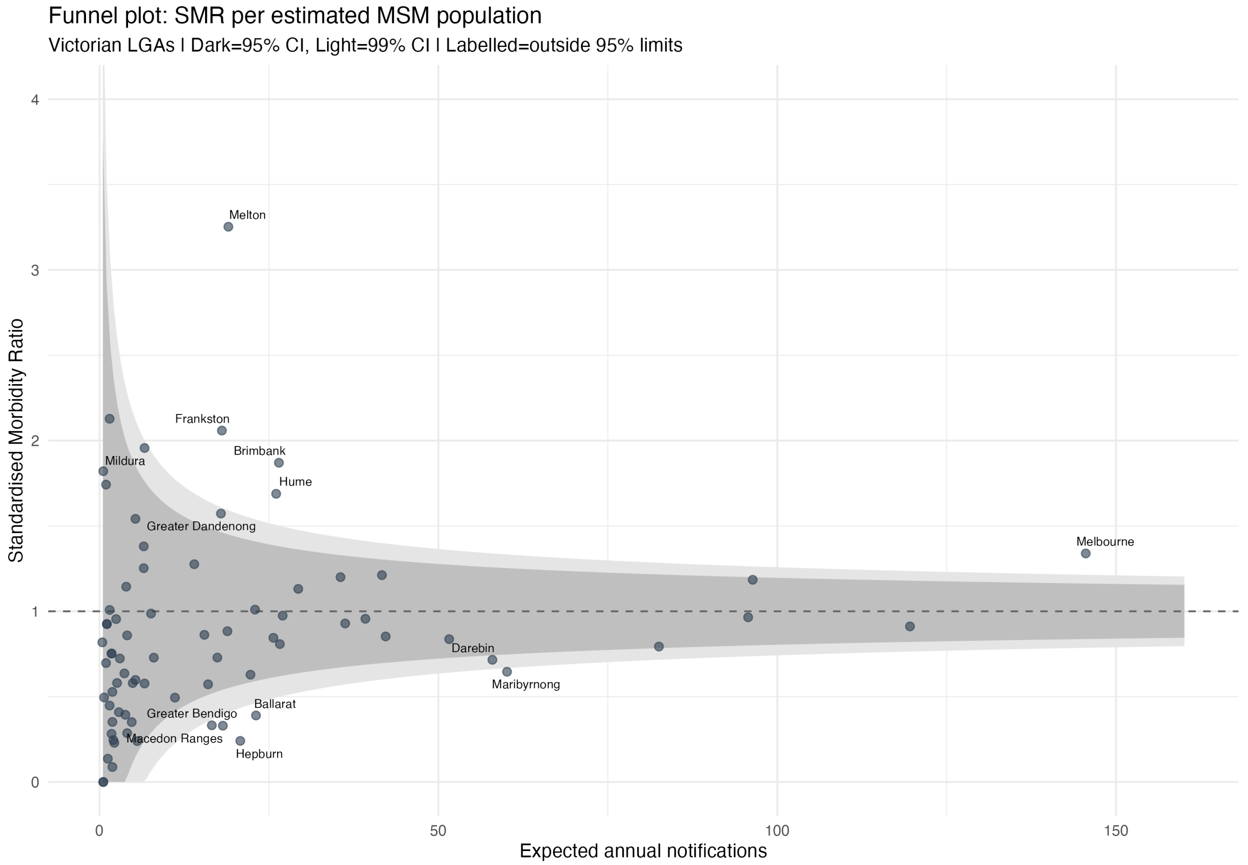

**Figure S8. Temporal stability: 2016 vs 2021 Census male-male couple counts by LGA.**

Each point represents one Victorian LGA. The near-perfect correlation (Pearson r = 0.983, Spearman rho = 0.972) demonstrates that the geographic distribution of same-sex male couples was highly stable across the 5-year inter-censal period, despite a 70% overall increase in counts (reflecting marriage equality legislation in December 2017 and increasing willingness to disclose). This supports the application of 2021 Census SSCF data as a proxy for MSM residential distribution during the 2019-2024 study period. Dashed line = line of equality; red line = linear fit.

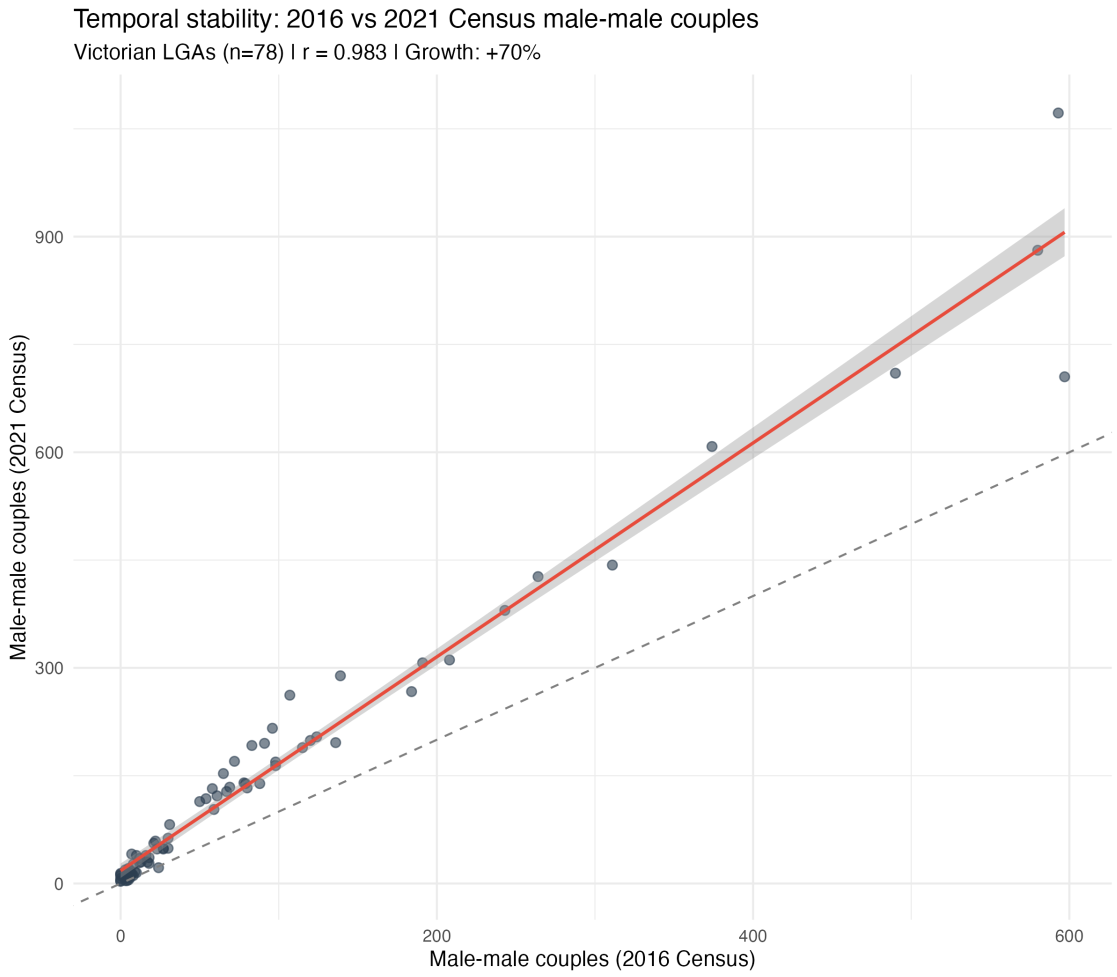
